# Menstrual Health in South India: Knowledge, Stigma, and Barriers to Participation

**DOI:** 10.64898/2026.09.16.26363278

**Authors:** Rupa Ravi, Joseph A. Amitrano, Dhruv R. Seshadri

## Abstract

Menstrual health remains a neglected public health challenge in India, where persistent stigma, incomplete education, inadequate sanitation, and limited access to menstrual health resources can adversely affect health and social participation. Institution-based educational programs represent an underexplored opportunity to improve menstrual health literacy while promoting more inclusive discussions surrounding menstruation. Bridging this gap, an exploratory, prospective educational evaluation was conducted at a hospital in Coimbatore, Tamil Nadu and an engineering college in Wayanad, Kerala. Anonymous pre- and post-course surveys assessed menstrual health knowledge, attitudes, practices, perceived barriers, and perspectives on emerging digital health technologies. Quantitative responses were summarized descriptively, and open-ended responses were reviewed to identify recurring perspectives that contextualized the quantitative findings. The standardized educational program integrated evidence-based menstrual health education with discussions of bioengineering and wearable technologies. Eighty-three participants completed the pre-course survey, 68 attended the educational program, and 20 completed the post-course survey. Because responses could not be linked at the individual level, the survey cohorts were compared descriptively. At baseline, 80 of 82 respondents (97.6%) recognized menstruation as a biological process, yet only 38 of 82 (46.3%) indicated that it was not solely a woman’s issue. The mean agreement that menstruation remained taboo was 7.44 of 10 (SD 1.99; n=78), and 51 of 77 respondents (66.2%) reported that they or others avoided activities during menstruation. Painful periods were identified as a barrier by 56 of 67 respondents (83.6%), and 18 of 67 (26.9%) considered institutional or community toilets insufficiently clean and private for menstrual management. Open-ended responses illustrated how pain, fear of leakage, stigma, and inadequate facilities constrained participation in school, physical activity, and social life. These findings document a marked gap between basic biological recognition and the social normalization and practical support of menstruation in two South Indian institutions, and support further evaluation of gender-inclusive, context-responsive menstrual health education using paired assessments, stronger follow-up, comparison groups, and long-term outcomes.

## 1. INTRODUCTION

Menstruation is a natural biological process experienced by approximately 1.8 billion girls, women, transgender men, and non-binary persons of reproductive age worldwide[1]. However, many menstruating individuals are unable to manage menstruation safely, comfortably, and with dignity. Menstrual health remains a neglected public health challenge in many low- and middle-income countries (LMICs), including India, where sociocultural stigma, incomplete menstrual health education, inadequate sanitation infrastructure, and limited access to affordable menstrual products continue to impede effective menstrual health management[2–5]. These barriers disproportionately affect individuals living in rural and underserved communities, where poverty, limited infrastructure, and restrictive social norms can compound existing health inequities[6,7]. Inadequate menstrual health support can affect educational participation, employment, psychosocial well-being, and gender equity[8]. Menstrual health equity is therefore relevant to achieving Sustainable Development Goal (SDG) 3 (Good Health and Well-being), SDG 5 (Gender Equality), and SDG 6 (Clean Water and Sanitation)[9–11]. Progress toward these goals requires access to menstrual products, comprehensive education, safe and private sanitation facilities, appropriate clinical support, and efforts to reduce the stigma that discourages individuals from seeking information and social support[2,12].

Although menstrual health indicators have improved nationally in India, substantial disparities persist. Analysis of the National Family Health Survey (NFHS-5) from more than 241,000 women aged 15–24 years demonstrated that the use of hygienic menstrual methods increased from 58% in 2015–2016 to 77% in 2019–2021[13]. However, this national gain was unevenly distributed: hygienic method use remained substantially lower among rural women (42.2%) than urban women (68.1%), lowest among tribal women (39%) compared with other groups, and ranged from only 19% among illiterate women to 69% among women with higher educational attainment, with a similar gradient by household wealth[14]. Cultural and religious taboos may further restrict participation in religious practices, food preparation, school, sports, and social activities while discouraging open discussion of menstrual and reproductive health[15–17]. A systematic review and meta-analysis of 138 studies involving more than 97,000 adolescent girls found that only 48% were aware of menstruation before menarche, 77% experienced cultural or religious restrictions during menstruation, 24% missed school during their menstrual period, and 37% were able to change menstrual absorbents at school[18]. These findings demonstrate that menstrual health is shaped by intersecting educational, social, economic, and infrastructural conditions.

Water, sanitation, and hygiene infrastructure is also closely associated with menstrual health. In a case-control study of 486 women in Odisha, India, the use of reusable menstrual absorbents was associated with greater odds of bacterial vaginosis, urinary tract infection, and self-reported symptoms of urogenital infection compared with the use of disposable sanitary pads[19]. Access to a private space for personal hygiene was associated with lower odds of bacterial vaginosis, emphasizing the importance of safe water and private washing and sanitation facilities[19]. Menstrual pain, fear of leakage, inadequate facilities, embarrassment, and social exclusion may also reduce school attendance, classroom concentration, participation in physical and social activities, and workforce productivity[20,21]. The nature and severity of these barriers vary considerably across geographic, socioeconomic, and cultural contexts, reinforcing the need for locally responsive interventions[7,8,22].

Tamil Nadu and Kerala provide important settings in which to examine these challenges. Tamil Nadu has made substantial progress in menstrual health, with NFHS-5 reporting that 98.3% of women aged 15–24 years used hygienic methods of menstrual protection[23]. Nevertheless, affordability, sanitation, menstrual health literacy, and access to appropriate products remain concerns, particularly in rural and lower-income communities. Kerala has similarly introduced initiatives such as the She Pad program to improve menstrual health support among school-aged girls[24]. However, disparities remain within rural districts such as Wayanad. Among 330 women tea plantation workers in Wayanad, 42.4% demonstrated satisfactory menstrual hygiene management, 10.6% reported difficulty affording menstrual products, 9.6% lacked appropriate toilet and washing facilities, and 16.1% reported at least one reproductive morbidity[25]. These findings indicate that strong statewide health indicators do not eliminate local educational, economic, and infrastructural barriers.

Educational interventions have improved menstrual health knowledge and practices among school-aged adolescents, but fewer studies have examined structured programs in higher-education and professional settings or programs involving both students and faculty members[26–29]. Existing interventions have also given limited attention to the combined roles of menstrual stigma, activity restrictions, institutional barriers, gender inclusion, and participant perspectives on emerging technologies[30]. Digital health tools, including smartphone applications and wearable devices, may offer additional opportunities for symptom tracking and individualized education, but they should complement rather than replace accessible health education, appropriate clinical care, adequate sanitation, and affordable menstrual products[31].

To address these gaps, our team developed and implemented a menstrual health educational program at two institutions in South India. The program combined evidence-based menstrual health education with discussions of bioengineering, wearable devices, and digital health technologies using the same standardized presentation and instructional approach at both sites.

This exploratory study evaluated menstrual health knowledge, attitudes, practices, perceived barriers, and perspectives on emerging technologies before and after the program. It also summarized open-ended responses concerning participants’ experiences of menstruation and menstrual health management. The findings were intended to inform the design and future evaluation of culturally responsive, gender-inclusive menstrual health education in resource-variable institutional settings.

## 2. METHODS

### 2.1 Study Setting

This prospective educational evaluation was conducted at two institutions in South India: 1) Healing Touch Rehabilitation Hospital in Coimbatore, Tamil Nadu and 2) Government Engineering College (GEC) Wayanad in Wayanad, Kerala. These sites were selected to evaluate the educational course across two distinct settings representing different healthcare, educational, and socioeconomic contexts within South India. Healing Touch Rehabilitation Hospital is a multidisciplinary rehabilitation center located in the metropolitan city of Coimbatore, which has comparatively greater access to healthcare services, menstrual health resources, and educational infrastructure. In contrast, GEC Wayanad is a public engineering institution located in the predominantly rural district of Wayanad, where disparities in menstrual health education, sanitation infrastructure, and access to menstrual hygiene resources remain more pronounced. Conducting the program at both institutions also allowed the study to explore perspectives on menstrual health management, stigma, sustainable menstrual products, and emerging digital health technologies across two institutional and geographical contexts.

### 2.2 Study Design and Educational Program

This prospective single-group educational evaluation used anonymous pre- and post-course surveys. Eighty-three participants completed the pre-course survey, 68 attended the educational program, and 20 completed the immediate post-course survey. Because the surveys did not include unique identifiers, pre- and post-course responses could not be linked at the individual level. The two survey cohorts were therefore compared descriptively rather than as paired observations. The same standardized presentation, educational content, and instructional approach were used at both institutions. The curriculum addressed menstrual physiology, common menstrual disorders, menstrual health management, menstrual products, cultural stigma, barriers in resource-variable settings, and the potential applications of bioengineering, wearable devices, and digital health technologies. Instruction combined didactic presentation, interactive discussion, examples of emerging health technologies, and opportunities for participant questions. The same learning objectives, presentation materials, and core discussion topics were used at both sites to maintain consistency in program delivery. The primary domains assessed were menstrual health knowledge, attitudes, practices, and perceived barriers. Participant perspectives on wearable technologies and other digital health innovations were evaluated as a secondary, exploratory domain. A detailed description of the standardized curriculum is provided (**Table 1**).

**Table 1.** Content and Delivery of the Standardized Menstrual Health Education Program.

| Program Characteristic | Healing Touch Rehabilitation Hospital | Government Engineering College |
| --- | --- | --- |
| Location | Coimbatore, Tamil Nadu | Wayanad, Kerala |
| Date | March 19, 2025 | March 21, 2025 |
| Participants | Rehabilitation hospital staff and healthcare-oriented participants | Engineering students and faculty members |
| Menstrual Health Content | Menstrual physiology; common menstrual disorders; menstrual health management; menstrual products; hygiene and infection prevention |  |
| Social and Public Health Content | Menstrual stigma and cultural perceptions; barriers to menstrual health management; challenges in resource-variable settings; effects on health and participation |  |
| Technology Content | Bioengineering approaches; wearable devices; remote monitoring; potential applications of digital health in menstrual and reproductive health |  |
| Instructional Format | Didactic presentation, interactive discussion, technology examples, and question-and-answer session |  |
| Presentation Materials | Same standardized presentation materials used at both sites |  |
| Pre-Course Assessment | Administered before the educational program |  |
| Post-Course Assessment | Distributed following the educational program |  |

### 2.3 Data Collection and Analysis

Paper-based questionnaires were administered immediately before the educational course. Following completion of the program, an electronic post-course survey was distributed using Google Forms (Google LLC, Mountain View, CA, USA). Survey responses were de-identified prior to analysis. Quantitative responses were summarized using descriptive statistics (Microsoft Excel, Version 16.111.2, Redmond, WA, USA). Pre- and post-course survey responses were compared descriptively across menstrual health knowledge, attitudes, practices, perceived barriers, and perspectives on digital health technologies. Wilson score 95% confidence intervals were calculated for selected proportions to demonstrate the precision of these descriptive estimates. Because the post-course survey represented a subset of attendees and responses were not paired at the individual level, paired statistical tests were not performed and comparisons were not interpreted as causal estimates of program effectiveness. Open-ended survey responses were reviewed and summarized to identify recurring perspectives concerning menstrual experiences, barriers to menstrual health management, effects on daily participation, stigma, and menstrual health technologies. These responses were used to provide context for the quantitative findings and were not analyzed to estimate the prevalence of individual experiences or themes. A geographic density map was generated to illustrate the regional distribution of participants across Tamil Nadu and Kerala (**Fig 1**).

**Figure 1.**
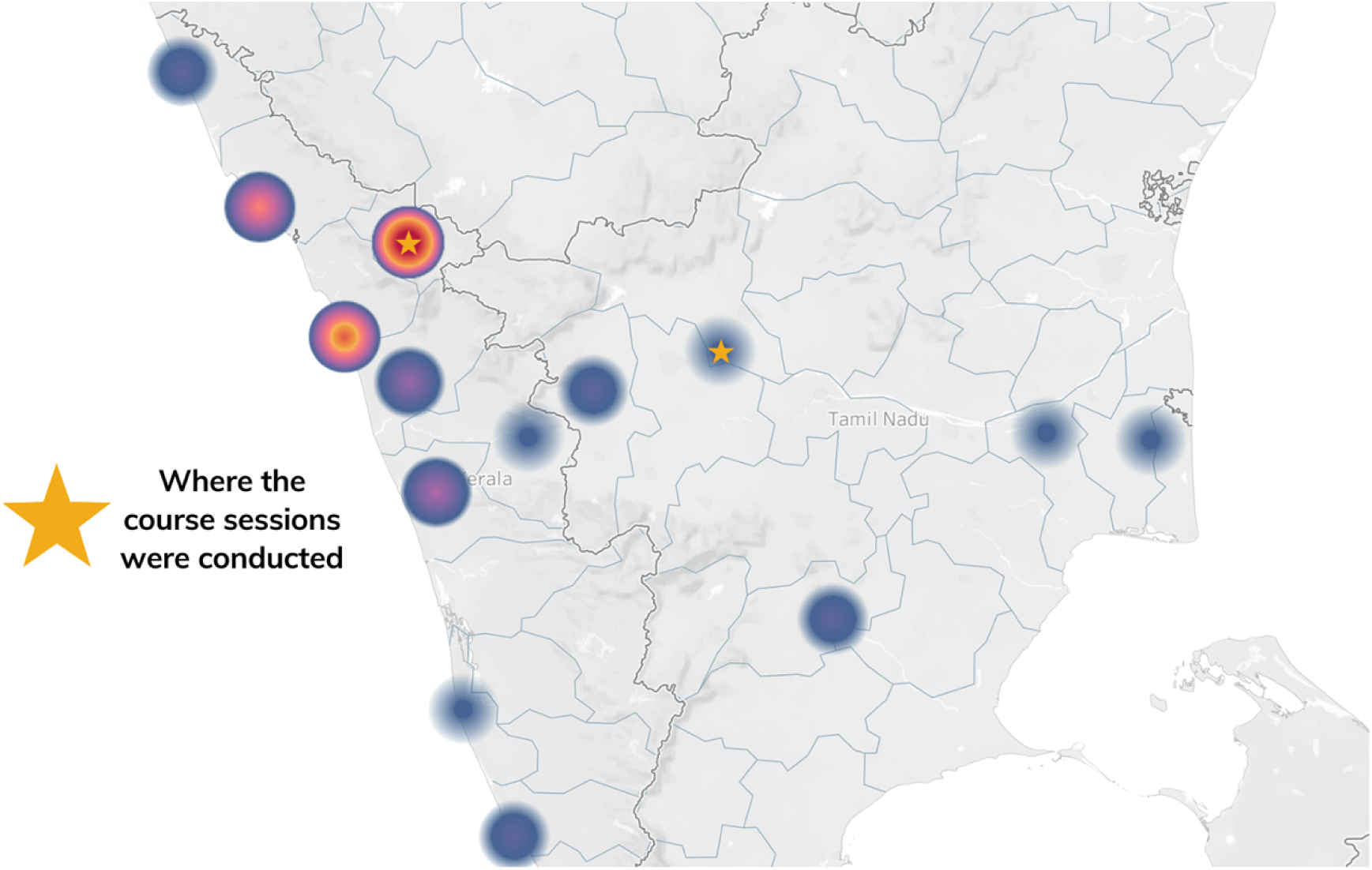
Density Map of Study Participant Distribution

**Figure 2.**
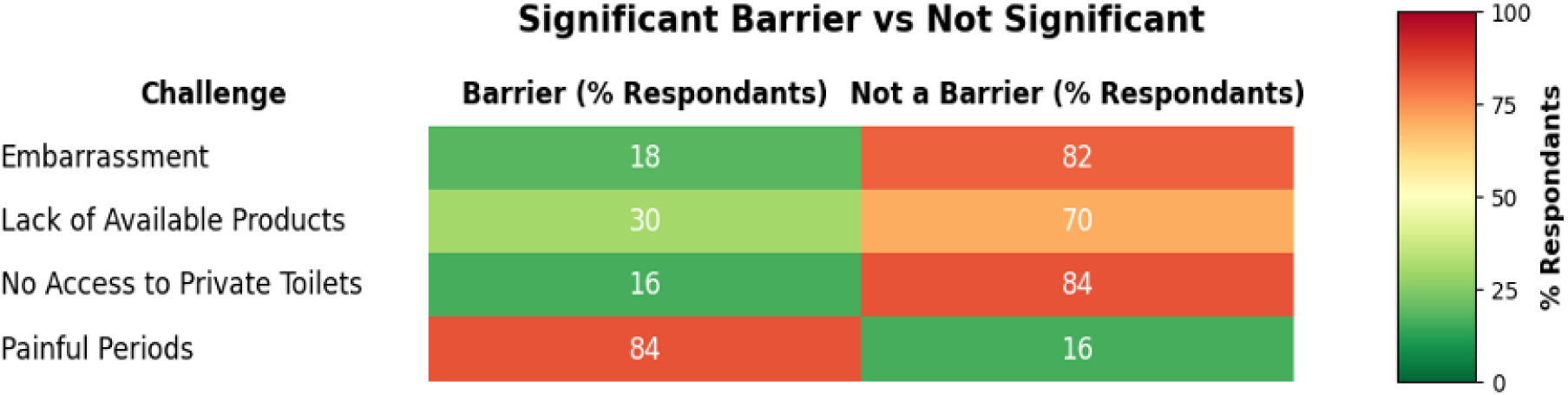
Heat Map of Menstrual Health Barriers Among Study Participants

### 2.4 Ethics

This study was reviewed by the Lehigh University Institutional Review Board through an administrative review process and was determined not to constitute human subjects research. All data analyzed in this study were fully de-identified, with no possibility of participant re-identification. Consequently, formal IRB approval and written informed consent for research were not required. Nevertheless, before electing to participate, all participants were informed of the purpose of the educational evaluation, the voluntary nature of participation, and their right to decline or withdraw without penalty.

## 3. RESULTS AND DISCUSSION

### 3.1 Participant Characteristics

A total of 83 participants completed the pre-course survey, 68 attended the educational program, and 20 completed the post-course survey. Among pre-course respondents, 70 (84.3%) were university students and 13 (15.7%) were faculty members or participants in other roles (**Table 2**). Most respondents identified as female (75/83, 90.4%), while eight (9.6%) identified as male. The inclusion of male participants was relevant to the program’s gender-inclusive approach; however, the small number of male respondents limited meaningful gender-stratified analysis. Malayalam was the most frequently reported language (89%), consistent with the large proportion of participants recruited at the Wayanad site, while Tamil (9.7%) and Hindi (1.3%) speakers represented participants from Coimbatore and other regions. Participants reported diverse religious backgrounds, including Hindu (54.2%), Muslim (24.1%), and Christian (18.1%) affiliations, with the remaining 3.6% reporting another affiliation. Household income was reported by only 23 participants. Among these respondents, 16 (69.6%) identified as middle-income, six (26.1%) as low-income, and one (4.3%) as high-income. The income distribution should be interpreted cautiously because data were missing for most participants.

**Table 2.** Participant Demographics.

| Characteristic | # of Respondents (n) | Percentage (%) |
| --- | --- | --- |
| Role |  |  |
| University Students | 70 | 84.3% |
| University Faculty | 13 | 15.7% |
| Gender |  |  |
| Female | 75 | 90.4% |
| Male | 8 | 9.6% |
| Primary Language Spoken at Home |  |  |
| Malayalam | 73 | 89.0% |
| Tamil | 8 | 9.8% |
| Hindi | 1 | 1.2% |
| Family Income Level (Self-Reported) |  |  |
| Low Income | 6 | 26.1% |
| Middle Income | 16 | 69.6% |
| High Income | 1 | 4.3% |
| Religion |  |  |
| Hindu | 45 | 54.2% |
| Muslim | 20 | 24.1% |
| Christian | 15 | 18.1% |
| Other | 3 | 3.6% |

The post-course cohort consisted of 20 respondents, including 16 students and four faculty members; all 20 identified as female. Consequently, the post-course cohort differed from the baseline cohort and did not include the male participants represented in the pre-course survey. Although the program reached participants from varied educational, linguistic, socioeconomic, and religious backgrounds, the convenience sample from two institutions should not be considered representative of university populations in South India.

### 3.2 Pre-Course Survey to Assess Menstrual Health Knowledge and Gendered Perceptions

The pre-course survey demonstrated high recognition of menstruation as a biological process alongside gaps in more specific menstrual health knowledge and varied perceptions of whether menstruation should be considered solely a women’s issue (**Table 3**). At baseline, 80 of 82 respondents (97.6%) identified menstruation as a biological process, and 67 of 82 (81.7%) correctly identified menstruation as the shedding of the uterine lining. Of the 82 respondents who answered the question concerning whether menstruation was solely a women’s issue, 39 (47.6%) responded that it was, 38 (46.3%) responded that it was not, and five (6.1%) were unsure.

**Table 3.** Pre-Course Knowledge, Attitudes, and Behaviors. Values are presented as n/N (%) unless otherwise indicated. Denominators vary because of missing, unsure, or non-codable responses. Gender-stratified estimates, particularly those for male participants, should be interpreted cautiously because of the small number of male respondents and item-level missingness. Menstrual-product items measured product awareness rather than personal product use.

| Category | Total (%) | Female (%) | Male (%) |
| --- | --- | --- | --- |
| **Knowledge** |  |  |  |
| Identified menstruation as a biological process | 80/82 (97.6%) | 74/74 (100%) | 6/8 (75.0%) |
| Identified menstruation as the shedding of uterine lining | 67/82 (81.7%) | 62/74 (83.8%) | 5/8 (62.5%) |
| Indicated that menstruation was not solely a woman's issue | 38/82 (46.3%) | 35/74 (47.3%) | 3/8 (37.5%) |
| Identified sanitary pads as a menstrual product | 81/81 (100%) | 74/74 (100%) | 7/7 (100%) |
| Identified menstrual cups as a menstrual product | 61/81 (75.3%) | 54/74 (73.0%) | 7/7 (100%) |
| Identified reusable cloth as a menstrual product | 49/81 (60.5%) | 46/74 (62.2%) | 3/7 (42.9%) |
| Identified tampons as a menstrual product | 42/81 (51.9%) | 39/74 (52.7%) | 3/7 (42.9%) |
| **Attitudes** |  |  |  |
| Agreement that menstruation remains a taboo topic, mean (SD) | 7.44 (1.99), n=78 | 7.66 (1.74), n=73 | 4.20 (2.77), n=5 |
| **Behaviors** |  |  |  |
| Reported activity avoidance during menstruation | 51/77 (66.2%) | 50/73 (68.5%) | 1/4 (25.0%) |

Among male respondents, five of eight (62.5%) indicated that menstruation was solely a women’s issue, compared with 34 of 74 female respondents with non-missing responses (45.9%). These subgroup findings should be interpreted cautiously because only eight male participants completed the baseline survey and no male participants completed the post-course survey. Nevertheless, the responses suggest that recognizing menstruation as a biological process does not necessarily translate into understanding menstrual health as a broader social and public health concern. This finding supports the inclusion of people of all genders in menstrual health education while underscoring the need for larger and more gender-diverse samples in future evaluations.

Open-ended responses illustrated the different ways participants interpreted the social relevance of menstruation. One male respondent who indicated that menstruation was not solely a women’s issue stated, *“Menstruation affects the mental state of women and indirectly that of the whole family.”* In contrast, one female respondent who characterized menstruation as a women’s issue explained, *“Because the one who gets periods and has to endure the pain are the women itself. Also the men don’t understand our discomfort.”* These responses suggest that participants’ views were shaped not only by biological knowledge but also by perceptions of pain, emotional effects, family relationships, and the extent to which non-menstruating individuals understand menstrual experiences.

Knowledge of menstrual products also varied. All 81 respondents (100%) identified sanitary pads as a product that could be used during menstruation, compared with 61 (75.3%) who identified menstrual cups, 49 (60.5%) who identified reusable cloth, and 42 (51.9%) who identified tampons. These findings reflect lower awareness of product options other than disposable sanitary pads, particularly tampons and reusable cloth. Menstrual health education may therefore benefit from presenting the range of available products and discussing their use, affordability, accessibility, acceptability, and safe maintenance without promoting a single product as appropriate for all individuals or settings. Collectively, the baseline findings indicate that menstrual health education in institutional settings should extend beyond basic menstrual physiology. A comprehensive approach should also address gendered perceptions, stigma, diverse menstrual experiences, and informed product choice. However, because the sample was recruited from two institutions and subgroup sizes were small, these findings should be interpreted as context-specific and hypothesis-generating.

### 3.3 Barriers to Menstrual Hygiene Management

Painful periods were the most frequently selected menstrual health barrier, reported by 56 of 67 respondents (83.6%). Other reported barriers included difficulty accessing menstrual products (20/67, 29.9%), embarrassment (12/67, 17.9%), and lack of private toilets (11/67, 16.4%). Open-ended responses described pain severe enough to limit participation in school, physical activity, travel, and social events. One participant reported, *“Severe pain and bleeding during first 3 days. Prefer not to participate in activities that needs physical strains. From 4th day onward, we don’t avoid anything due to menstruation.”* This response illustrates how menstrual pain and heavy bleeding could substantially affect daily functioning rather than represent a minor inconvenience. The prominence of pain in this study is consistent with previous research identifying dysmenorrhea as an important contributor to reduced educational participation and quality of life[20,21].

Agreement that menstruation remained a taboo topic was high at baseline, with a mean rating of 7.44 out of 10 (SD 1.99; n=78). Lack of education was identified as a contributor to menstrual shame or embarrassment by 60 of 81 respondents (74.1%), cultural or religious beliefs by 49 of 81 (60.5%), and fear of discussing private matters by 48 of 81 (59.3%). These findings suggest that menstrual stigma was shaped by intersecting educational, cultural, and interpersonal factors. Such stigma may influence whether individuals feel comfortable discussing menstruation, seeking information or care, obtaining menstrual products, and participating in everyday activities during menstruation [15–18]. These social barriers were accompanied by changes in daily participation. Fifty-one of 77 respondents (66.2%) reported that they or others avoided at least one activity during menstruation, including school, sports, travel, social events, entering the kitchen, or consuming certain foods. Open-ended responses attributed avoidance to pain, fatigue, fear of leakage or staining, embarrassment, and culturally based restrictions. These findings suggest that menstruation-related activity avoidance may arise from the combined effects of physical symptoms, stigma, concerns about menstrual leakage, and social expectations rather than from a single factor.

Sanitation conditions presented an additional barrier. Among 67 respondents who evaluated institutional or community toilets, 40 (59.7%) reported that the facilities were sufficiently clean and private for menstrual management, 18 (26.9%) reported that they were not, and nine (13.4%) were unsure. Thus, approximately two in five respondents either lacked confidence in the adequacy of available facilities or considered them inadequate. Limited access to clean toilets, private washing spaces, and appropriate disposal systems may make menstrual management more difficult and discourage participation in educational and community activities [17,19].

Collectively, these findings indicate that menstrual health management was shaped by interacting physical, educational, social, and infrastructural factors. Education alone may be insufficient if participants continue to experience severe menstrual symptoms, stigma, inadequate sanitation, or limited access to products. Comprehensive approaches should therefore combine menstrual health education with appropriate clinical referral pathways, supportive social environments, access to suitable products, and menstrual-supportive water, sanitation, and hygiene infrastructure.

### 3.4 Descriptive Pre- and Post-Course Differences in Menstrual Health Knowledge and Attitudes

Selected knowledge, attitude, and behavior responses differed descriptively between the pre- and post-course cohorts (**Fig 3**). Correct identification of menstruation as a biological process was reported by 80 of 82 pre-course respondents (97.6%; 95% CI 91.5%-99.3%) and 20 of 20 post-course respondents (100%; 95% CI 83.9%-100%). Correct identification of menstruation as shedding of the uterine lining was reported by 67 of 82 respondents before the program (81.7%; 95% CI 72.0%-88.6%) and 18 of 20 afterward (90.0%; 95% CI 69.9%-97.2%). Recognition that menstruation was not solely a woman’s issue was reported by 38 of 82 respondents before the program (46.3%; 95% CI 36.0%-57.1%) and 10 of 19 afterward (52.6%; 95% CI 31.7%-72.7%).

**Figure 3.**
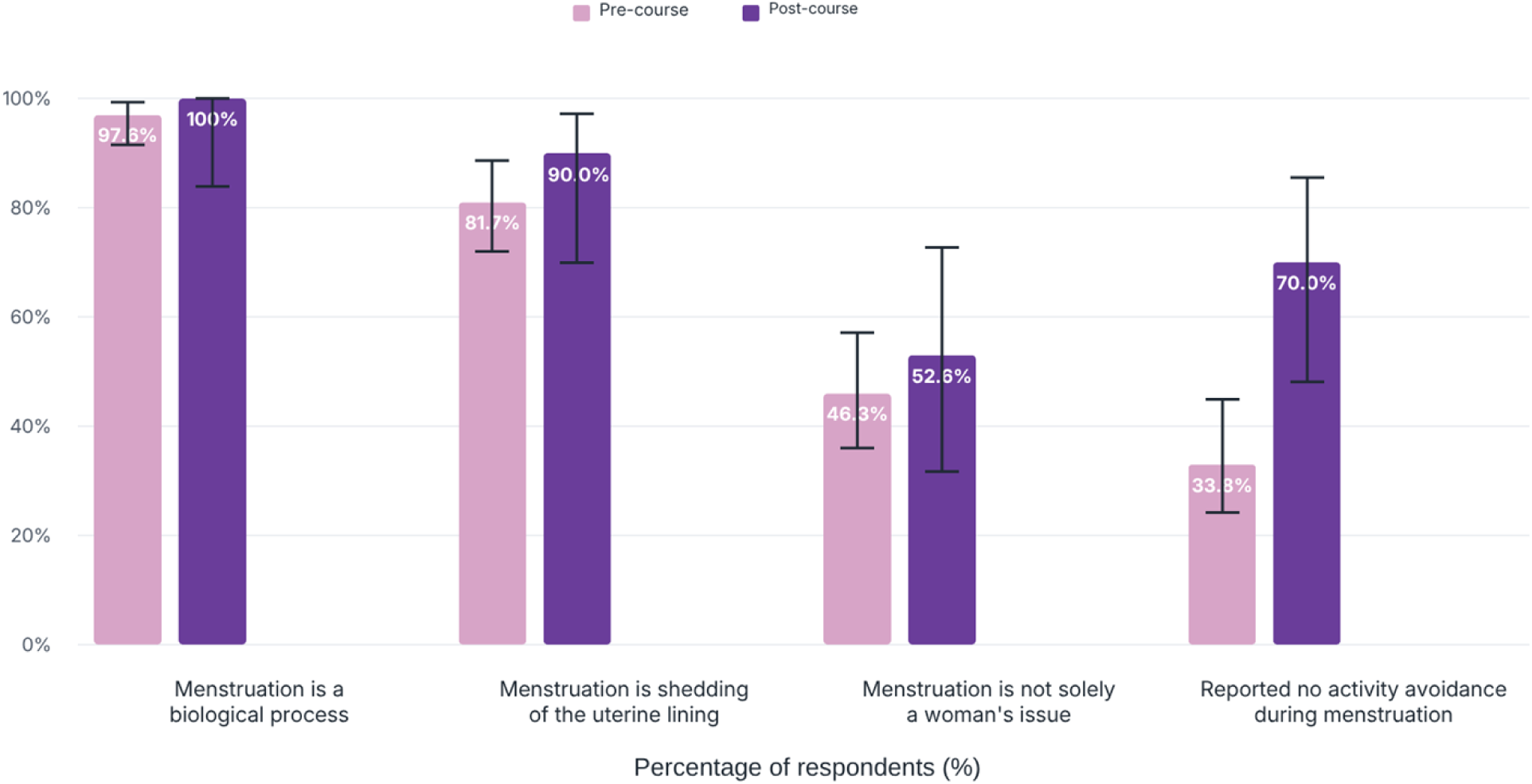
Selected Responses in the Pre- and Post-Course Survey Cohorts. Bars represent the percentage of respondents providing each response, and error bars represent Wilson 95% confidence intervals. Pre- and post-course surveys were anonymous and could not be linked at the individual level; therefore, the figure presents descriptive comparisons between separate respondent cohorts rather than within-person changes.

The wide confidence intervals, particularly for the post-course estimates, demonstrate the imprecision associated with the small post-course sample and preclude firm conclusions about between-cohort differences.

Mean agreement that menstruation remained a taboo topic was 7.44 out of 10 before the program (SD 1.99; n=78) and 6.70 afterward (SD 2.05; n=20). Because this item used a 1-10 scale, it was summarized as a continuous measure rather than converted into a percentage. Activity avoidance was reported by 51 of 77 pre-course respondents (66.2%) and 6 of 20 post-course respondents (30.0%). Conversely, 26 of 77 respondents before the program reported no activity avoidance (33.8%; 95% CI 24.2%-44.9%), compared with 14 of 20 afterward (70.0%; 95% CI 48.1%-85.5%). Although this difference was notable, the surveys did not establish whether the same participants changed their behavior, and the finding may reflect differences between the two respondent groups.

Post-course respondents rated the educational program favorably, with a mean rating of 4.45 out of 5 (SD 0.69; n=20). Open-ended feedback emphasized the importance of education in addressing menstrual stigma and promoting community awareness. One respondent stated, *“We need to change societal views about menstruation. Having periods is not a bad thing.”* This statement illustrates a post-course perspective that framed menstruation as a normal aspect of health and emphasized the need for broader social change. Additionally, among the 18 participants who answered the advocacy item, 11 (61.1%) rated their willingness to advocate for improved menstrual health within their communities as four or five on a five-point scale. This finding suggests that some respondents perceived a potential role for themselves in community engagement and menstrual health awareness. Overall, the observed patterns were consistent with the possibility that structured menstrual health education may support knowledge, discussion, and more inclusive attitudes. However, only 20 of the 68 course attendees completed the post-course survey, corresponding to a post-course response rate of 29.4%, and all post-course respondents were female. Because responses were anonymous and unpaired, the observed differences may reflect respondent selection, social desirability, pre-existing differences between the cohorts, or exposure to the educational program. These findings should therefore be considered exploratory and hypothesis-generating. Future evaluations should use linked assessments, stronger follow-up procedures, comparison groups, and longer-term measurement to determine whether educational programs produce sustained changes in knowledge, attitudes, behavior, and community engagement.

### 3.5 Participant Perspectives on Digital Health

Digital health and bioengineering were included as secondary, exploratory components of the educational program. Among respondents who provided a codable answer to the post-course question about valuable learning, 67% referred to technological advances in women’s health, biosensors, biomedical devices, or related innovations. These responses suggest that some participants valued the interdisciplinary content; however, they should not be interpreted as evidence of technology access, adoption, usability, clinical effectiveness, or likely future use. Digital health tools, including smartphone applications, wearable sensors, and other biomedical devices, may support menstrual health education, symptom tracking, and communication with healthcare professionals. However, this study did not evaluate participants’ access to technology, digital literacy, willingness to use specific tools, data privacy concerns, clinical validity, or health outcomes. Digital health technologies should therefore be considered potential complements to menstrual health education, menstrual-supportive WASH infrastructure, affordable products, and appropriate clinical care rather than substitutes for these foundational resources. Future studies should examine the accessibility, acceptability, affordability, privacy, and clinical value of specific technologies in resource-variable settings.

### 3.6 Public Health Implications and Future Directions

The findings suggest that improving menstrual health requires more than increasing access to menstrual products. Despite high baseline recognition of menstruation as a biological process, participants reported gaps in menstrual health knowledge, substantial stigma, activity restrictions, menstrual pain, and concerns about sanitation infrastructure. The observed pre- and post-course differences were encouraging but cannot be attributed to the educational program because the surveys were unpaired and the post-course response rate was low. Nevertheless, the findings indicate that menstrual health initiatives should combine comprehensive education with access to appropriate clinical care, menstrual-supportive water, sanitation, and hygiene infrastructure, affordable products, and community engagement. Educational programs may help normalize discussion of menstruation and address misconceptions, but education alone is unlikely to overcome structural barriers such as inadequate sanitation, limited product access, or insufficient clinical support for severe menstrual symptoms. Higher-education and professional training institutions may provide valuable settings for menstrual health education, given documented gaps in university students’ menstrual health experiences and evidence that structured educational interventions improve outcomes [28,29]. Incorporating discussion of sustainable menstrual products, bioengineering, and digital health may encourage interdisciplinary engagement with menstrual health challenges. However, technology-focused content should remain grounded in community needs and should not displace attention from sanitation, affordability, clinical care, cultural acceptability, accessibility, and data privacy.

Prior Indian educational interventions have largely focused on adolescent girls in school settings and primarily assessed knowledge, attitudes, hygiene practices, and broader sexual and reproductive health outcomes (**Table 4**) [32–36]. In contrast, the present study evaluated an interdisciplinary program in higher-education and professional institutional settings, included female and male participants in the baseline cohort, and considered pain, stigma, activity participation, sanitation, sustainability, and emerging health technologies together. These features broaden the dimensions examined but do not offset the limitations of the small, unpaired post-course cohort.

**Table 4.** Selected educational interventions addressing menstrual health in India.

| Location | Population | Sample size | Setting | Study design | Intervention approach | Outcomes assessed | Follow-up | Key finding | Ref |
| --- | --- | --- | --- | --- | --- | --- | --- | --- | --- |
| Maharashtra | Adolescent girls, grades 8-10 | 250 | 3 rural schools | Single-group pre/post intervention | Didactic education on menstrual hygiene, practices, health risks, and taboos | Knowledge, attitudes, and practices | 2 months | Awareness of hygienic practices before menarche was 34.8% at baseline; significant practice changes included handwashing after pad changes ($p=0.05$). | [32] |
| Odisha | Girls in grades 9-10 | 790 baseline; 760 end-line | 8 government girls' high schools | Cluster randomized trial | Comprehensive school-based sexual and reproductive health education using presentations and handbooks | Knowledge, attitudes, and practices across puberty, menstruation, pregnancy, contraception, and infections | 3 months | Recognition of normal pubertal changes increased from 60.1% to 94.8%; positive perception of menstruation increased from 63.3% to 93.5% in the intervention arm. | [33] |
| Karnataka | Menstruating female high-school students | 585 | High schools in and around BG Nagara | Single-group pre/post intervention | Pharmacist-mediated menstrual hygiene education | Menstrual knowledge, attitudes, and hygiene practices | Post-intervention | At baseline, 43.8% had good menstrual knowledge and 86.0% used sanitary pads; knowledge and practice scores improved significantly after education ($p<0.001$). | [34] |
| Gujarat | Adolescent girls aged 12-16 years | 100 (50 per group) | Adolescent educational setting | Two-group quasi-experimental study | Eight-week skills-based education versus lecture-based education | Knowledge, attitudes, and practices | Immediate and 2 months | Skills-based education produced greater immediate gains in knowledge ($p<0.001$), attitudes ($p<0.001$), and practices ($p=0.006$); gains persisted at 2 months. | [35] |
| Karnataka | Adolescent girls | 67 | 2 schools | Pre/post pilot evaluation | Validated PALMS module covering puberty, anatomy, menstruation, hygiene, myths, symptoms, and self-care | Module feasibility, menstrual-health knowledge, and participant feedback | Immediate | The module was pilot-tested with 67 students; feasibility and pre/post knowledge change were assessed to support larger-scale evaluation. | [36] |
| Tamil Nadu and Kerala | University students, faculty, and healthcare-oriented participants; female and male participants | 83 pre-course; 68 attendees; 20 post-course | Engineering college and rehabilitation hospital | Exploratory prospective evaluation; unpaired pre/post cohorts | Standardized interdisciplinary education on physiology, disorders, products, pain, stigma, participation, WASH, sustainability, and emerging health technologies | Knowledge, attitudes, stigma, practices, activity participation, barriers, sanitation, and technology perspectives | Immediate | At baseline, 83.6% identified pain as a barrier and 66.2% reported activity avoidance; no activity avoidance was 33.8% pre-course versus 70.0% post-course (descriptive, unpaired cohorts). | Present study |

Future multicenter studies should include larger and more gender-diverse samples, linked pre- and post-course assessments, stronger follow-up procedures, and appropriate comparison groups. Longer-term assessments are also needed to determine whether educational programs influence knowledge retention, stigma, healthcare-seeking behavior, activity participation, advocacy, or menstrual health outcomes. Future evaluations should additionally examine which program components are most useful across different institutional and cultural contexts.

## 4. STRENGTHS AND LIMITATIONS

A principal strength of this study was its implementation across two distinct institutional settings in South India, including a rehabilitation hospital and an engineering college. The evaluation examined menstrual health knowledge alongside physical, social, behavioral, and infrastructural barriers, providing a broader assessment than knowledge-focused evaluations alone. The program included students, faculty members, and participants of different genders and incorporated sustainable menstrual products, gender inclusion, bioengineering, and digital health as interdisciplinary topics. Open-ended responses also provided contextual information about participants’ experiences and perspectives.

Several limitations substantially constrain interpretation. First, only 20 of the 68 course attendees completed the post-course survey, corresponding to a response rate of 29.4% and creating substantial potential for nonresponse and selection bias. Reasons for nonresponse were not systematically documented. Second, anonymous responses could not be linked at the individual level; therefore, the findings represent comparisons between separate pre- and post-course cohorts rather than within-person changes. The post-course cohort also differed demographically from the baseline cohort and consisted entirely of female respondents. Third, the study did not include a control or comparison group. A future cluster-randomized evaluation could compare institutions receiving the program immediately with those receiving standard coursework without the added menstrual health programming, before later receiving the program themselves, while collecting the same measures at baseline and follow-up. Such a design would better distinguish program-associated changes from other baseline biases. In the present study, observed differences may therefore reflect respondent selection, testing effects, social desirability, or pre-existing differences between the cohorts rather than effects of the educational program. Fourth, outcomes were self-reported and assessed immediately after the program, preventing evaluation of longer-term knowledge retention, behavioral change, advocacy, or health outcomes. Fifth, participants were recruited from only two institutions, limiting generalizability to other institutions, regions, and populations. Although the same standardized curriculum was delivered at both sites, differences in institutional setting and participant composition may have influenced responses. The sample was insufficient to evaluate site-specific differences. Finally, only eight male participants completed the baseline survey, and no male participants completed the post-course survey, limiting conclusions regarding gender-specific perspectives or the effects of gender-inclusive education. In light of these limitations, the findings should be considered exploratory and hypothesis-generating.

## 5. CONCLUSION

This exploratory study documented intersecting menstrual health barriers among students, faculty members, and other participants at two institutions in South India. Its clearest finding was a gap between biological recognition and lived social conditions, where 97.6% recognized menstruation as a biological process, yet only 46.3% indicated that it was not solely a woman’s issue, the mean taboo rating was 7.44 of 10, and 66.2% reported activity avoidance. Participants also identified menstrual pain and concerns about sanitation infrastructure. Descriptive differences between the pre- and post-course cohorts were encouraging but cannot establish program effectiveness because responses were unpaired and post-course participation was limited. The findings reinforce the need for menstrual health strategies that combine evidence-based education with stigma reduction, menstrual-supportive water, sanitation, and hygiene infrastructure, affordable products, appropriate clinical care, and community engagement. Educational institutions may provide useful settings for interdisciplinary menstrual health education, while digital health and bioengineering technologies may complement, but should not replace, these foundational resources. Larger studies using linked assessments, comparison groups, more gender-diverse samples, and longer follow-up are needed to evaluate whether such programs produce sustained changes in knowledge, attitudes, behavior, and health outcomes. These efforts may support the integration of comprehensive menstrual health education into higher education institutions as an approach for advancing the Sustainable Development Goals related to health, education, gender equality, and sanitation while promoting menstrual equity both within India and other resource-variable settings.

### Author Reflexivity Statement

This study was conceived, designed, implemented, and analyzed by investigators from Lehigh University as part of an educational initiative focused on menstrual health in South India. Healing Touch Rehabilitation Hospital (Coimbatore, Tamil Nadu) and Government Engineering College Wayanad (Kerala) served as host institutions, providing access to participants and facilities that enabled delivery of the educational course and data collection. The study was conducted with the support and permission of institutional leadership at both sites, and the educational content was developed for the local context and delivered consistently at both sites. The authors recognize the importance of conducting global health research through respectful engagement with local institutions and communities and acknowledge the contributions of the participating institutions and course attendees in making this work possible.

## Data Availability

Data is available from the authors upon reasonable request.

## Acknowledgements

The authors thank the faculty, students, and staff of Healing Touch Rehabilitation Hospital (Coimbatore, Tamil Nadu) and Government Engineering College Wayanad (Kerala) for their participation and support in facilitating this educational program. The authors also acknowledge the institutional leadership at both sites for providing the facilities and logistical support necessary to conduct the menstrual health education course. Most importantly, the authors thank all study participants for their time, engagement, and willingness to share their experiences and perspectives on menstrual health.

## Author Contributions

**Conceptualization:** Rupa Ravi, Dhruv R. Seshadri

**Methodology:** Rupa Ravi, Dhruv R. Seshadri

**Investigation:** Rupa Ravi, Dhruv R. Seshadri

**Data Curation:** Rupa Ravi

**Formal Analysis:** Rupa Ravi

**Visualization:** Rupa Ravi, Joseph Amitrano

**Writing – Original Draft:** Rupa Ravi, Joseph Amitrano, Dhruv R. Seshadri

**Writing – Review & Editing:** Rupa Ravi, Joseph Amitrano, Dhruv R. Seshadri

**Supervision:** Dhruv R. Seshadri

**Project Administration:** Rupa Ravi, Dhruv R. Seshadri

**Funding Acquisition:** Dhruv R. Seshadri

